# Clinical advancement of patients infected with SARS-CoV-2 Omicron variant in Shanghai, China

**DOI:** 10.1101/2022.07.03.22277169

**Authors:** Jiasheng Shao, Rong Fan, Tiejun Zhang, Catherine Lee, Xuyuan Huang, Fei Wang, Haiying Liang, Ye Jin, Ying Jiang, Yanhua Gu, Jianrong Hu, Gang Huang

**Affiliations:** Department of Infectious Diseases and Immunology, Jiading District Central Hospital, Shanghai University of Medicine and Health Sciences, Shanghai 201899, China; Genomics, Center for Molecular and Cellular Bioengineering, Biotechnology Center, Technische Universität Dresden 01307, Germany; Department of Epidemiology, School of Public Health, Fudan University, Shanghai 200032, China; College of Public Health, University of Georgia, Athens, GA 30602, USA; Department of Urology, Renji Hospital, Shanghai Jiaotong University, Shanghai 200127, China; Department of Intensive Care Unit, Jiading District Central Hospital, Shanghai 201899, China; Department of Respiratory Medicine, Jiading District Central Hospital, Shanghai 201899, China; Department of Nursing, Jiading District Central Hospital, Shanghai 201899, China; Shanghai Key Laboratory of Molecular Imaging, Shanghai University of Medicine and Health Sciences, Shanghai 201318, China

**Keywords:** SARS-CoV-2, Omicron variant, Vaccination, PCR conversion, Paxlovid

## Abstract

**Background:** Studies on the Omicron variant infection have generally been restricted to descriptions of its initial clinical and epidemiological characteristics. We investigated the timeline-related advancement in individuals with Omicron variant.

**Methods:** We conducted a retrospective, single-centered study including 226 laboratory-confirmed cases with Omicron variant between April 6th and May 11th, 2022 in Shanghai, China. Final date of follow-up was May 30, 2022.

**Results:** Among 226 enrolled patients, the median age was 52 years old, and 118 (52.2%) were female. The duration from onset of symptoms to hospitalization was 3 days (interquartile range [IQR]: 2-4 days) for symptomatic patients. Cough occurred in 168 patients (74.3%). The median interval to negative reverse-transcriptase PCR tests of nasopharynx swab was 10 days ([IQR]: 8-13 days). No radiographic progressions were found in 196 patients on the 7th day after onset of symptoms. The median duration of fever in all participants was 5 days (IQR: 4–6 days). The median PCR conversion time of Paxlovid-treated patients was 8 days (IQR: 7-10 days) compared with that of Lianhuaqingwen (10 days, IQR: 8-13 days) (p=0.00056). Booster vaccination can significantly decrease the severity of Omicron infection when compared with unvaccinated, partially or fully vaccinated patients (p=0.009). The median time of PCR conversion in individuals aged under 14 recovered significantly faster than older patients (median 8 days versus 11 days, P < 0.0001). In multivariate logistic analysis, erythrocyte sedimentation rate (ESR) (OR=1.05) was independently related to the severity of the infection.

**Conclusions:** The majority clinical symptoms of Omicron infection were not severe. Early and aggressive administration of Paxlovid can significantly reduce the PCR conversion time. Booster vaccination should also be highly recommended in population over 14 years old.

## Introduction

The novel Omicron variant was first reported to the World Health Organization (WHO) by Gauteng province, South Africa in mid-November last year, and shortly after was classified as a variant of concern (VOC)(1). The novel variant shares several mutations with the previous VOC Alpha, Beta, and Gamma variants, which spread over more than 100 countries and immediately lifted up international concerns due to its higher transmissibility and infectivity(2, 3).

Clinical characteristics of the Omicron variant have been partially described. Kim et al. reported 40 cases and Lee et al. reported 83 cases of SARS-CoV-2 Omicron in South Korea, respectively, suggesting the patients in their studies had no clinical symptoms or mild symptoms and had recovered within several days(4, 5). Another report with 43 patients recruited from Center for Disease Control and Prevention (CDC) of the U.S. indicated that the most commonly reported symptoms were cough, fatigue, and rhinorrhea. Only one vaccinated individual was hospitalized for 2 days, and no deaths were reported in their study(6).

Since August 2021, China adopted a dynamic zero-COVID strategy to respond to SARS-CoV-2 variants with higher transmissibility and successfully stemmed hundreds of COVID-19 outbreaks that are associated with imported cases(7). Even under local administration’s aggressive containment, the variant spread very fast across Shanghai since mid-March 2022, and arrived at its peak on April 13, 2022, with about 25,000 infected individuals in one single day (**Figure 1**). As of May 11, 2022, 617,979 people in Shanghai had been infected(8). However, the clinical course of the Omicron is still not fully interpreted. Herein, we collected data from 226 laboratory-confirmed cases in our hospital, as one of 44 designated hospitals for treating SARS-CoV-2 Omicron in Shanghai, aiming to present the clinical progress and outcome of Omicron variant infection.

**Fig. 1.**
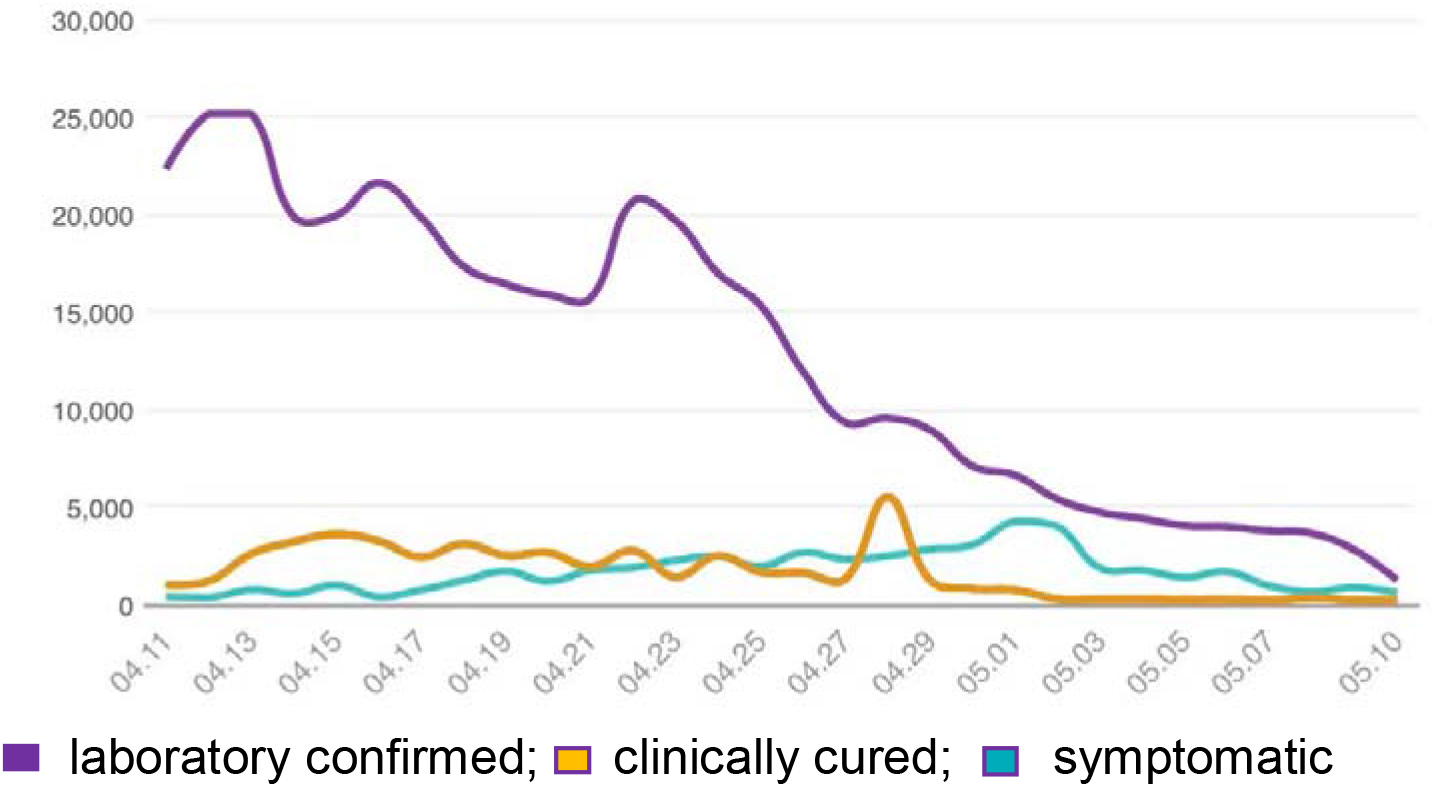
Tendency map of laboratory confirmed, clinically cured and symptomatic cases infected with Covid-19 Omicron variant from April 11 to May 10, 2022

## Methods

### Study designs and participants

For this retrospective, single-center study, we recruited patients from April 18, 2022 to May 11, 2020, at Jiading District Central Hospital, Shanghai, China, an officially-designated hospital that treats patients with the Omicron variant. Symptomatic patients with the Omicron variant in this study were enrolled from mobile cabin hospitals, fever clinics and nursing homes. There were four asymptomatic patients also enrolled because of unstable conditions such as dizziness and sudden elevation of blood pressure. Criteria for admission, diagnoses, therapy and discharge were made based on the most updated version of the national COVID-19 protocol (9). This study was approved by the Ethics Committee of Jiading District Central Hospital and informed consent was obtained from all patients involved before data was collected. Informed consent was waived for patients who were unable to obtain an informed consent.

### Procedures

Information of demographic, epidemiological, clinical symptoms and signs, underlying co-existing disorders, laboratory and radiological findings, treatment and outcomes were collected from medical records and radiological images were scrutinized by a well-trained team of physicians. In clinical settings, the date of onset of disease was defined as the day when the symptom(s) was found. Negative PCR conversion of the upper respiratory tract was defined as an interval between the first date of PCR positivity and the first date of two consecutively negative days by testing nasopharyngeal swab. Body temperature was measured by nurses using mercury thermometers at least four times a day. Fever was defined as an axillary temperature of 37.5°C or higher. The definition of defervescence was if the body temperature could be maintained normal without administration of any antipyretics.

### Laboratory confirmation

Laboratory confirmation of the Omicron variant infection was accomplished by the Shanghai Center for Disease Prevention and Control (CDC). Subsequent test of nasopharyngeal swab specimen for the variant after hospitalization was performed by both Jiading District Central Hospital and Shanghai CDC based on the recommendation by the National Institute for Viral Disease Control and Prevention (China).

### Statistical analysis

Means and interquartile range (IQR) for continuous variables were compared via independent group *t* tests. Frequencies and proportions for categorical variables were compared with the chi-square test. The outcome of this study was measured as PCR conversion days. The outcomes were compared within different possible affecting variables by Kaplan–Meier curves. The association between baseline clinical characteristics, laboratory findings and factors associated with the severity of infection were calculated with logistical regression. All statistical analyses were performed using SAS software version 9.4 (SAS institute, Cary, NC, USA). A p value < 0.05 was considered statistically significant.

## Results

### Clinical and laboratory profiles of participants on admission

As of May 11, 2022, a total of 226 patients were enrolled in this study. The demographic distribution of Omicron variant infected people in Shanghai was shown in **Figure 2**. The median age was 52 years old (IQR, 32–68 years) and 118 (52.2%) were female. 92(40.7%) patients were unvaccinated, 7(3.1%), 56(24.8%), and 71 (31.4%) patients received 1 dose (partially), 2 doses(fully), and 3 doses(booster) vaccination, respectively, with CoronaVac (SARS-CoV-2 Inactivated Vaccine [Vero Cell]) manufactured by SINOVAC BIOTECH CO.,LTD. The median duration from onset of symptoms to hospital admission was 3 (2–3) days in patients with symptoms. 105 patients (46.5%) had one or more chronic comorbidities. Hypertension was the most common coexisting disorder (76 [33.6%]), followed by diabetic mellitus (30[13.3%]). Most common symptoms on admission are cough (168 [74.3%]), sputum production (111 [49.1%]), and fever (103 [45.6%]). Less common symptoms included sore throat, fatigue, dizziness and headache, rhinorrhea, loss of olfaction, myalgia/arthralgia, diarrhea and shortness of breath (**Table 1**).

**Figure 2.**
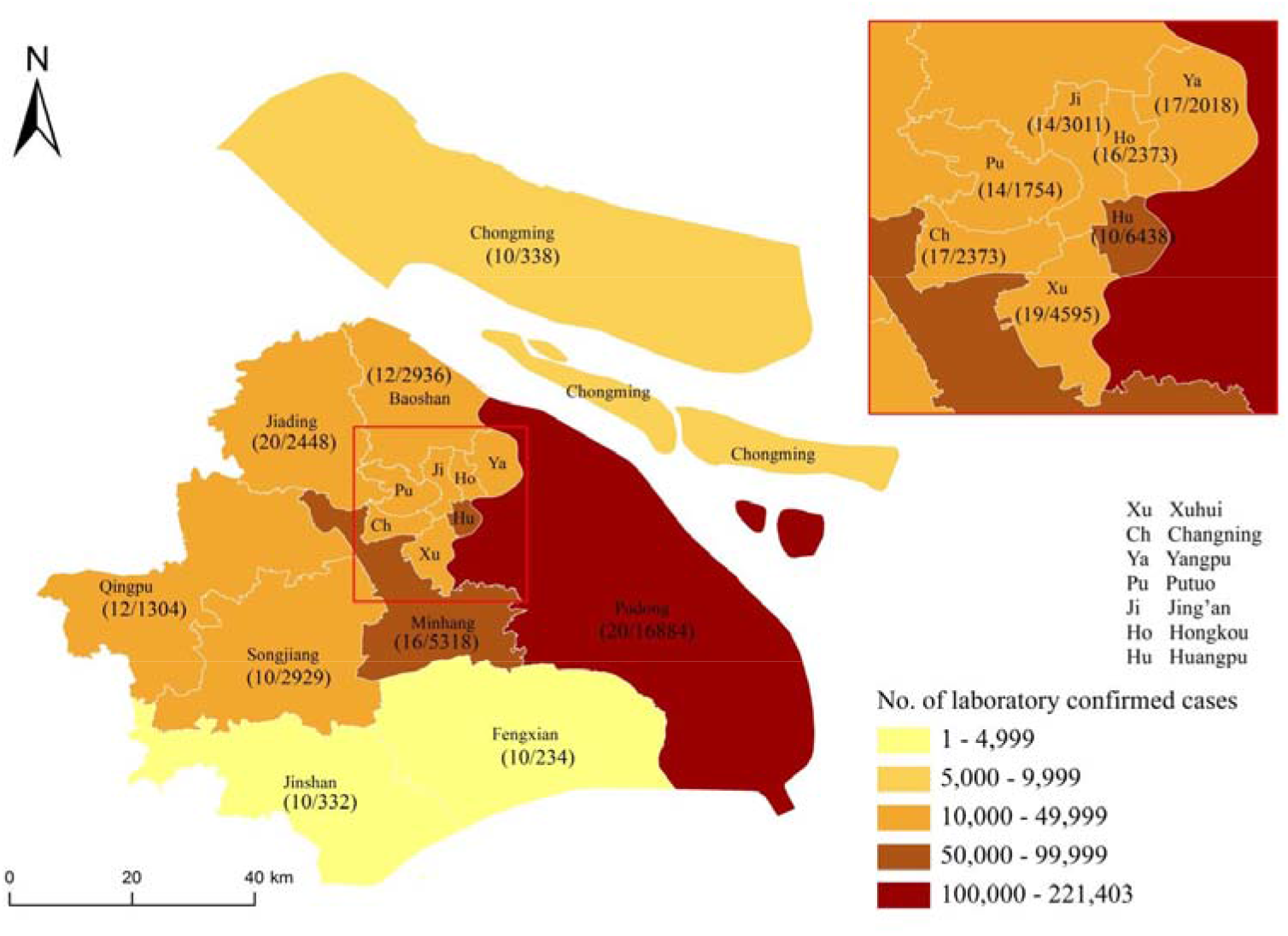
Distribution of patients with the Omicron variant across Shanghai from March 1 to May 11, 2022 Shown are the official data of all documented, laboratory-confirmed cases of the Omicron variant throughout Shanghai, according to the Shanghai Municipal Health Commission as of May 11, 2022. The numerator indicates the number of patients who were included in the cohort study and the denominator indicates the number of cases with clinical symptoms for each district, as reported by the Shanghai Municipal Health Commission

**Table 1.** Clinical characteristics of patients infected with Omicron variant on admission (N=226).

| characteristic | All patients<br>(N=226) |  | Disease Severity<sup>a</sup> |  |  |  |
| --- | --- | --- | --- | --- | --- | --- |
|  |  | Asymptomatic<br>(N=4) | Mild<br>(N=180) | Moderate<br>(N=41) | Severe<br>(N=1) | p value |
| Age |  |  |  |  |  |  |
| Median (IQR) - yr. | 52.0(32.0-68.0) | 51.5(43.5-58.5) | 48.0(28.5-62.5) | 71.0(57.0-78.0) | 75s | <0.001 |
| Distribution —<br>no./total no. (%) |  |  |  |  |  | <0.001 |
| 0–14 yr. | 28(12.4) | 0(0.0) | 28(15.6) | 0(0.0) | 0 |  |
| 15–49 yr. | 73(32.4) | 1(25.0) | 66(36.9) | 6(14.6) | 0 |  |
| 50–64 yr. | 52(23.1) | 2(50.0) | 42(23.5) | 8(19.5) | 0 |  |
| ≥65 yr. | 72(32.0) | 1(25.0) | 43(24.0) | 27(65.9) | 1(100) |  |
| Female sex —<br>no./total no. (%) | 118(52.2) | 2(50.0) | 99(55.0) | 17(41.5) | 0 | 0.251 |
| Smoking history —<br>no./total no. (%) |  |  |  |  |  | 0.245 |
| Smoker | 27(11.9) | 0(0.0) | 21(11.7) | 6(14.3) |  |  |
| Non-smoker | 199(88.1) | 4(100.0) | 159(88.3) | 36(85.7) |  |  |
| Median days from<br>one set of<br>symptoms to<br>admission | 3.0(2.0-3.0) | / | 3.0(2.0-3.0) | 3.0(2.0-4.0) | 2.0(2.0-2.0) | 0.608 |
| Median times of<br>vaccinations(IQR)-<br>dose | 2.0(0.0-3.0) | 3.0(2.0-3.0) | 2.0(0.0-3.0) | 0.0(0.0-2.0) | 0.0(0.0-0.0) | 0.009 |
| Fever— no./total<br>no. (%) | 103(45.6) | 0(0.0) | 83(46.1) | 20(48.8) | 0(0.0) | 0.226 |
| Median<br>temperature (IQR)<br>— °C | 38.4(38.0-39.0) | / | 38.4(38.0-39.0) | 38.4(37.9-38.8) |  | 0.444 |
| Duration of fever | 5.0(4.0-6.0) | / | 5.0(4.0-6.0) | 6.5(6.0-8.3) | 14.0(12.0-16.0) | <0.001 |
| Distribution of<br>Temperature —<br>no./total no. (%) |  |  |  |  |  | 0.280 |
| <37.5°C | 123(54.4) | 4(100.0) | 97(53.9) | 21(51.2) | 1(100.0) |  |
| 37.5–38.0°C | 35(15.5) | 0(0.0) | 28(15.6) | 7(17.1) | 0(0.0) |  |
| **38.1–39.0°C** | 51(22.6) | 0(0.0) | 38(21.1) | 13(31.7) | 0(0.0) |  |
| **>39.0°C** | 17(7.52) | 0(0.0) | 17(9.4) | 0(0.0) | 0(0.0) |  |
| **Symptoms — no. (%)** |  |  |  |  |  |  |
| **Cough** | 168(74.3) | 0(0.0) | 133(73.9) | 34(82.9) | 1(100.0) | **0.005** |
| **Sputum production** | 111(49.1) | 0(0.0) | 85(47.2) | 25(61.0) | 1(100.0) | **0.031** |
| **Loss of olfaction** | 16(7.1) | 0(0.0) | 14(7.8) | 2(4.9) | 0(0.0) | 0.822 |
| **Fatigue** | 33(14.6) | 0(0.0) | 27(15.0) | 6(14.6) | 0(0.0) | 1.000 |
| **Dizziness and Headache** | 28(12.4) | 0(0.0) | 22(12.2) | 6(14.6) | 0(0.0) | 0.801 |
| **Shortness of Breath** | 4(1.8) | 0(0.0) | 3(1.67) | 1(2.44) | 0(0.0) | 0.600 |
| **Rhinorrhea** | 18(8.0) | 0(0.0) | 16(8.89) | 2(4.88) | 0(0.0) | 0.696 |
| **Sore throat** | 44(19.5) | 0(0.0) | 40(22.2) | 4(9.76) | 0(0.0) | 0.212 |
| **Diarrhea** | 6(2.7) | 0(0.0) | 3(1.67) | 3(7.32) | 0(0.0) | 0.196 |
| **Myalgia/arthralgia** | 11(4.9) | 0(0.0) | 8(4.44) | 3(7.32) | 0(0.0) | 0.559 |
| **Comorbidity — no. (%)** |  |  |  |  |  |  |
| **Respiratory system diseases** | 6(2.7) | 0(0.0) | 4(2.2) | 2(4.9) | 0(0.0) | 0.396 |
| **Diabetic mellitus** | 30(13.3) | 0(0.0) | 18(10.0) | 12(29.3) | 0(0.0) | **0.012** |
| **Hypertension** | 76(33.6) | 1(25.0) | 50(27.8) | 24(58.5) | 1(100.0) | **<0.001** |
| **Non-hypertension cardiovascular disease** | 13(5.8) | 0(0.0) | 10(5.6) | 3(7.3) | 0(0.0) | 0.787 |
| **Cerebrovascular disease** | 22(9.7) | 0(0.0) | 9(5.00) | 13(31.7) | 0(0.0) | **<0.001** |
| **Hepatitis B virus infection** | 1(0.4) | 0(0.0) | 1(0.6) | 0(0.0) | 0(0.0) | 1.000 |
| **Cancer** | 14(6.2) | 0(0.0) | 6(3.3) | 7(17.1) | 1(100.0) | **0.001** |
| **Chronic renal disease** | 2(0.9) | 0(0.0) | 2(1.1) | 0(0.0) | 0(0.0) | 1.000 |
| **Endocrine disorder** | 2(0.9) | 0(0.0) | 2(1.1) | 0(0.0) | 0(0.0) | 1.000 |
| **asthma** | 2(0.9) | 0(0.0) | 1(0.6) | 1(2.4) | 0(0.0) | 1.000 |
| **Parkinson disease** | 2(0.9) | 0(0.0) | 1(0.6) | 1(2.4) | 0(0.0) | 0.366 |
| **immunodeficiency** | 1(0.4) | 0(0.0) | 0(0.0) | 0(0.0) | 1(100.0) | 0.204 |

<sup>a</sup> Asymptomatic: Omicron variant nucleic acid positive confirmed by Reverse Transcription Polymerase Chain Reaction (RT-PCR) without clinical symptoms;
Mild: mild clinical manifestations such as fever, cough, expectoration, fatigue, etc.; without radiologic signs of pneumonia; Moderate: clinical signs mentioned above; with radiologic signs of pneumonia; Severe: clinical manifestations mentioned above, with radiologic signs of pneumonia, plus anyone of the following: 1) Shortness of breath, respiration rate (RR) ≥ 30 times/min; 2) In the state of resting and inhalation of air, the oxygen saturation(SaO2) is ≤93%; 3) PaO2/FiO2 ≤300mmHg (1mmHg=0.133kpa); PaO2/FiO2 shall be corrected according to the following formula in high altitude areas (more than 1000m above sea level): PaO2 / FiO2 × [760/atmospheric pressure (mmHg)]; 4) Progressive aggravation of clinical symptoms mentioned above, and chest computered tomography (CT) showed significant progression of lesions > 50% within 24~ 48 hours; Critical: 1) Respiratory failure occurs and mechanical ventilation is required; 2) Shock; 3) Patients with other organ failure need ICU monitoring and treatment.

### Regimen and clinical outcomes of the study population

All patients were admitted to isolation wards. Patients were given supportive treatment after admission. An antiviral drug-Paxlovid was used in 17 patients (13.0%) to assess its effectiveness and Lianhuaqingwen capsules (LHQW) were used in 114 patients above 14 years old, respectively. The median interval of PCR conversion for Paxlovid-treated patients was 8 days (IQR: 7-10 days) compared to that in LHQW-treated patients (10 days, IQR: 8-13days) (p=0.00056) (**Fig 3)**. 42 patients received antibacterial therapy (moxifloxacin, 18 [42.9%]; ceftriaxone, 24 [57.1%]). To the time of submission, a total of 224 (99.1%) patients were discharged after 14 (12–17) days hospitalization and 2 patients were still in intensive care unit (ICU). No patients perished.

**Fig. 3.**
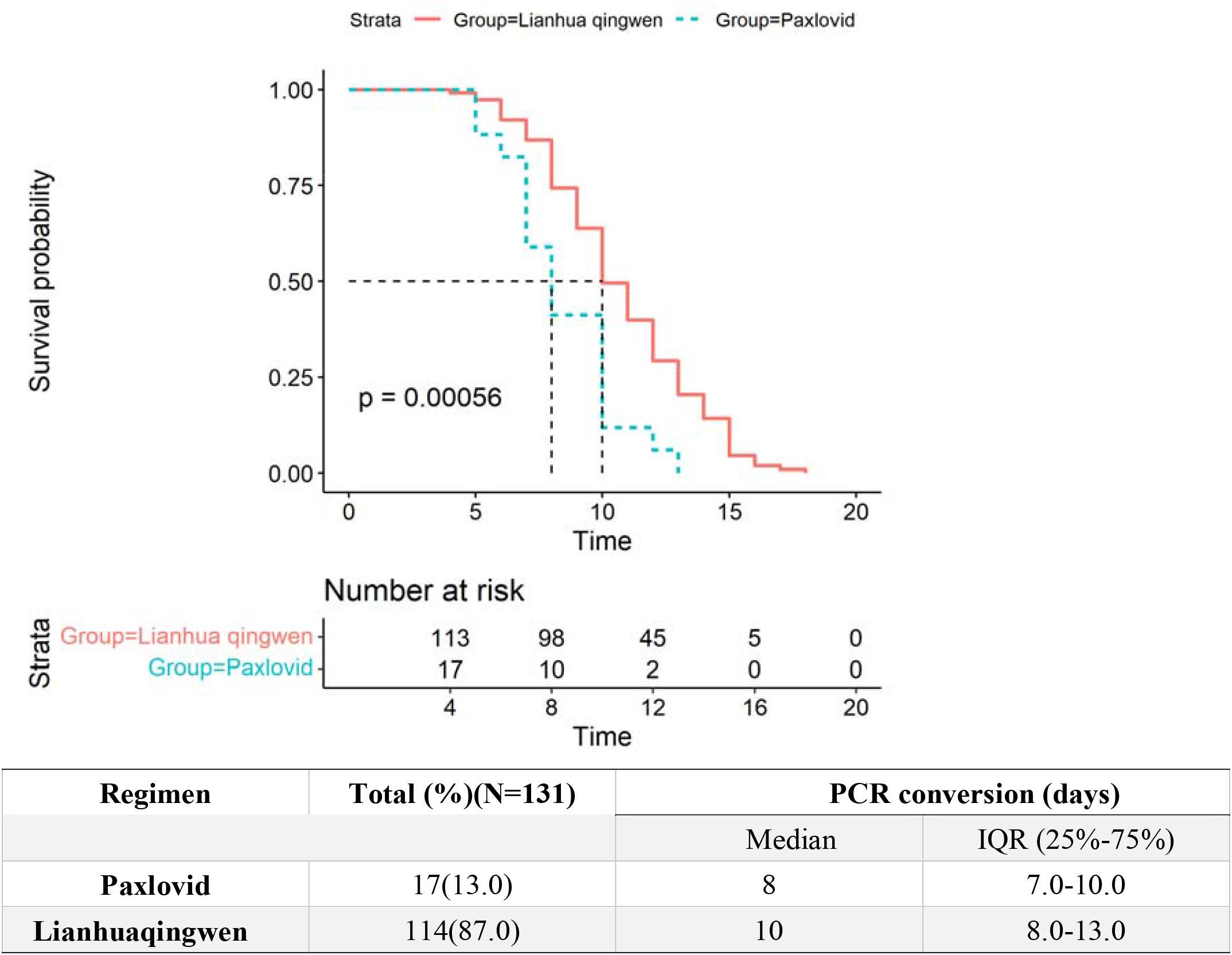
The comparison of PCR conversion time between Paxlovid- and Lianhuaqingwen-treated patients.

### Duration of fever in the study population

As fever was one of the cardinal signs among patients with Covid-19(10), we investigated the defervescence period in this population. Fever occurred in 103(45.6%) patients during their clinical course. Body temperature gradually got back to normal with or without supportive therapy after hospitalization. The median duration of fever in all participants was 5 days (IQR: 4–6 days) after onset of symptoms. The median duration of fever in moderate patients was 6.5 days (IQR: 6.0–8.3 days). Two severe cases including a male patient in his 90s who developed from moderate to severe had significantly longer duration of fever (14 days, IQR: 12-16 days) (**Fig.4)**. Most other symptoms including cough, expectoration and sore throat, fatigue, dizziness and headache also relinquished before fever abatement.

**Fig 4.**
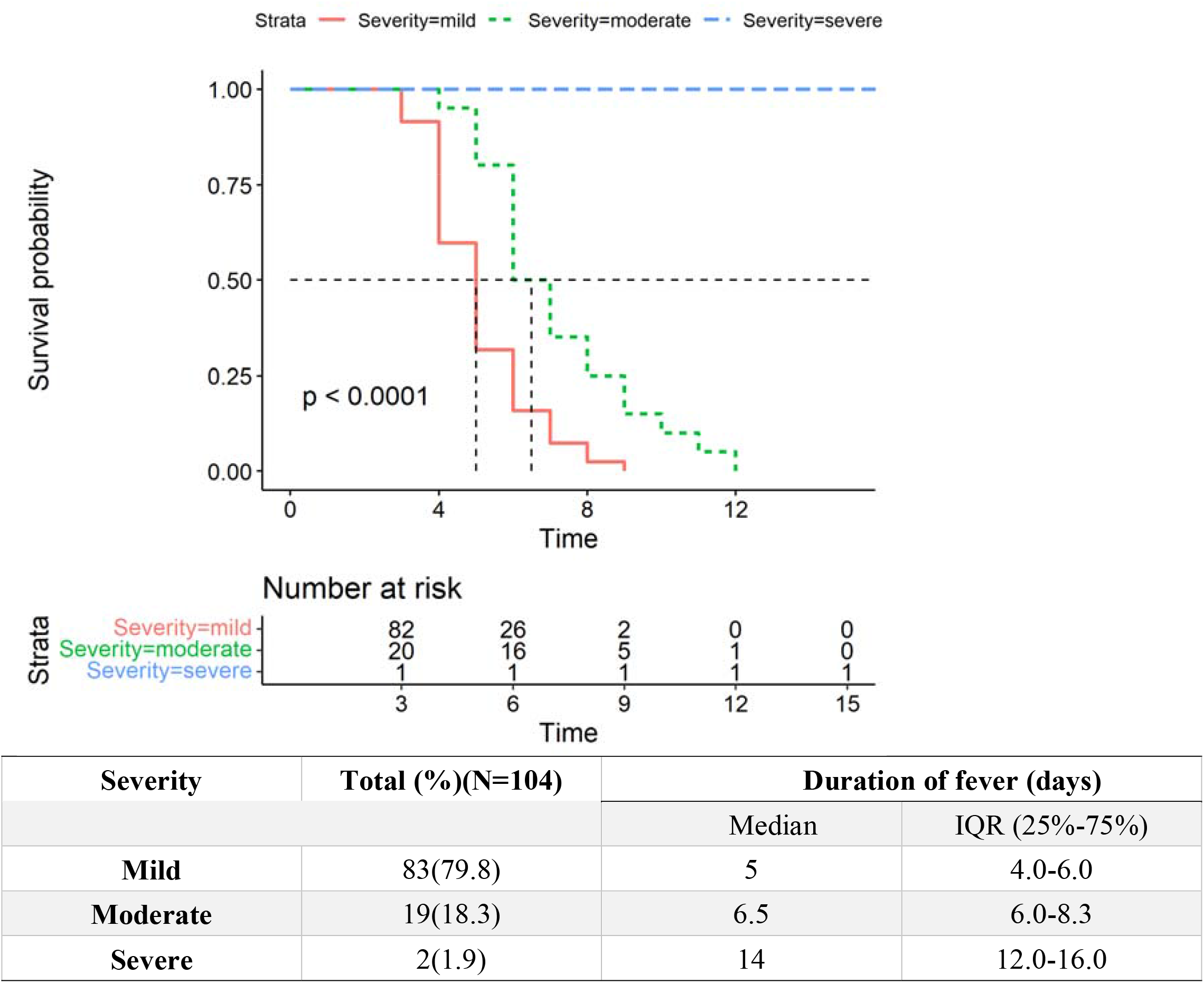
Duration of fever in mild, moderate and severe cases.

### Imaging changes in disease progression

Chest computed tomography (CT) or X-ray were performed for 198 patients excluding 28 children upon admission, 11 patients showed unilateral lesions, 31 patients were manifested with bilateral lesions while the remaining 156 adult patients were normal. Radiological examinations were repeated in 198 patients, no progressions were shown in 196 cases (99.0%) after 7 days of symptoms onset. Only two senile male patients deteriorated in imaging. The representative dynamic imaging changes of three patients with different severities were shown in **Fig 5**.

**Figure 5.**
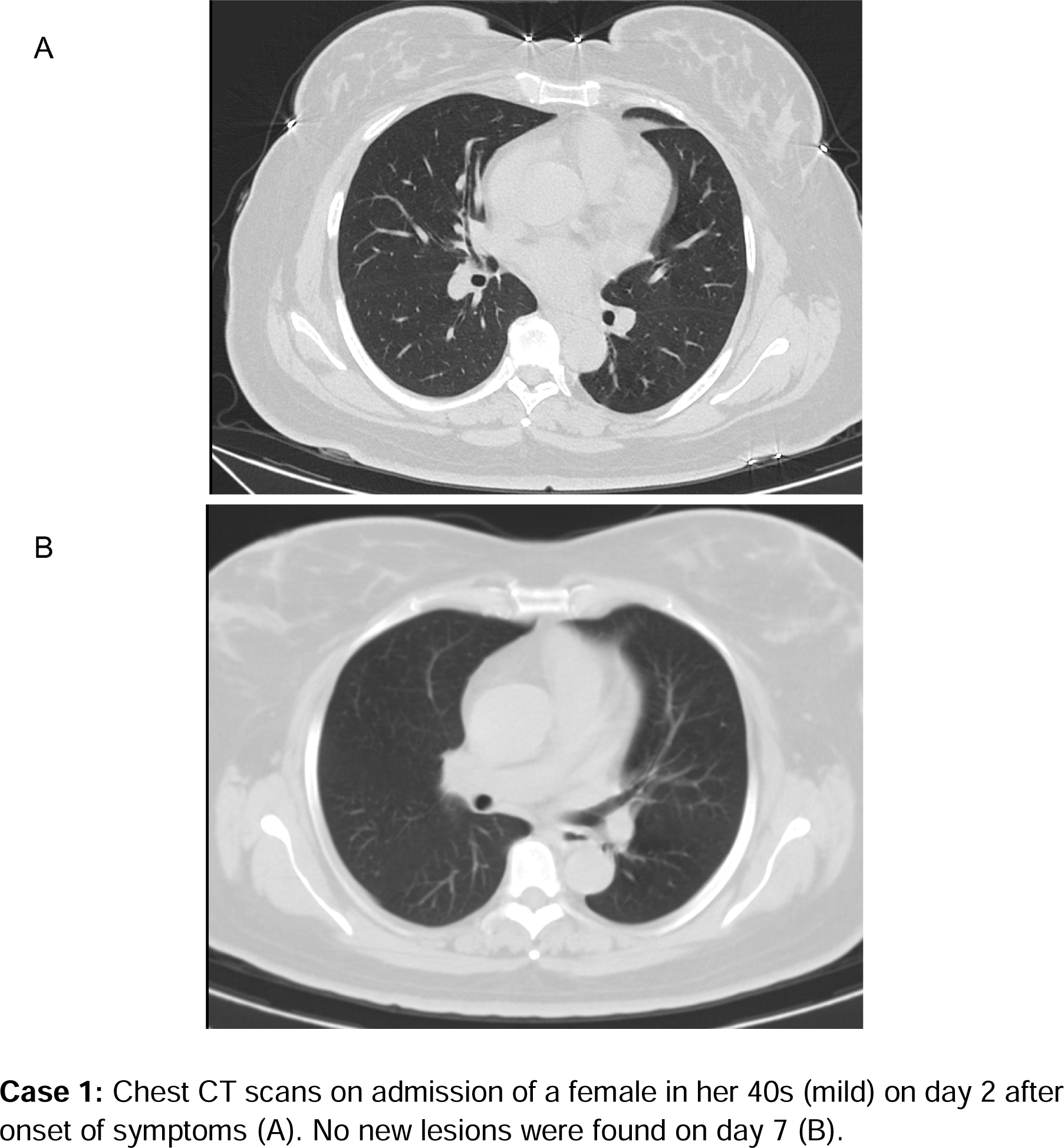
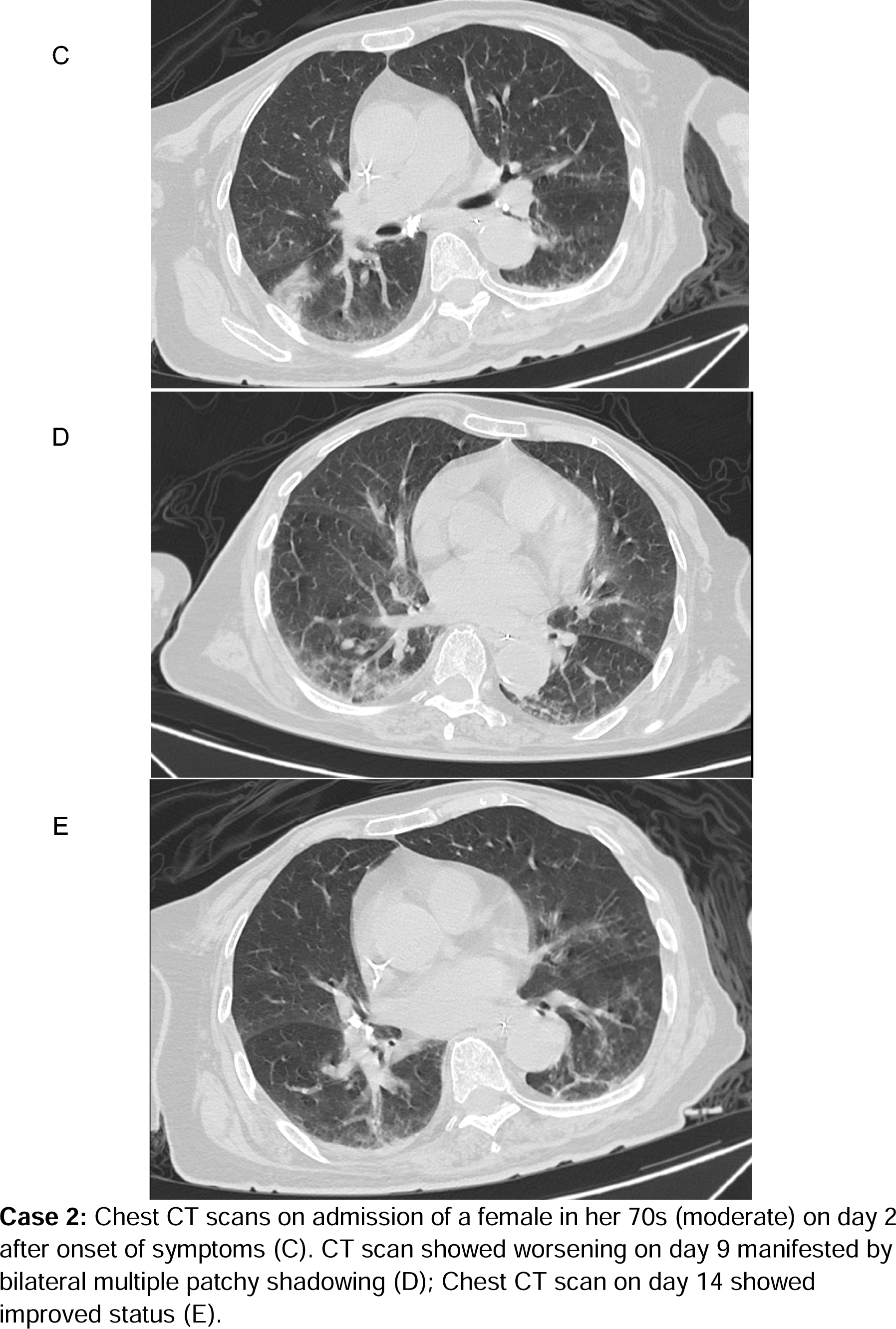
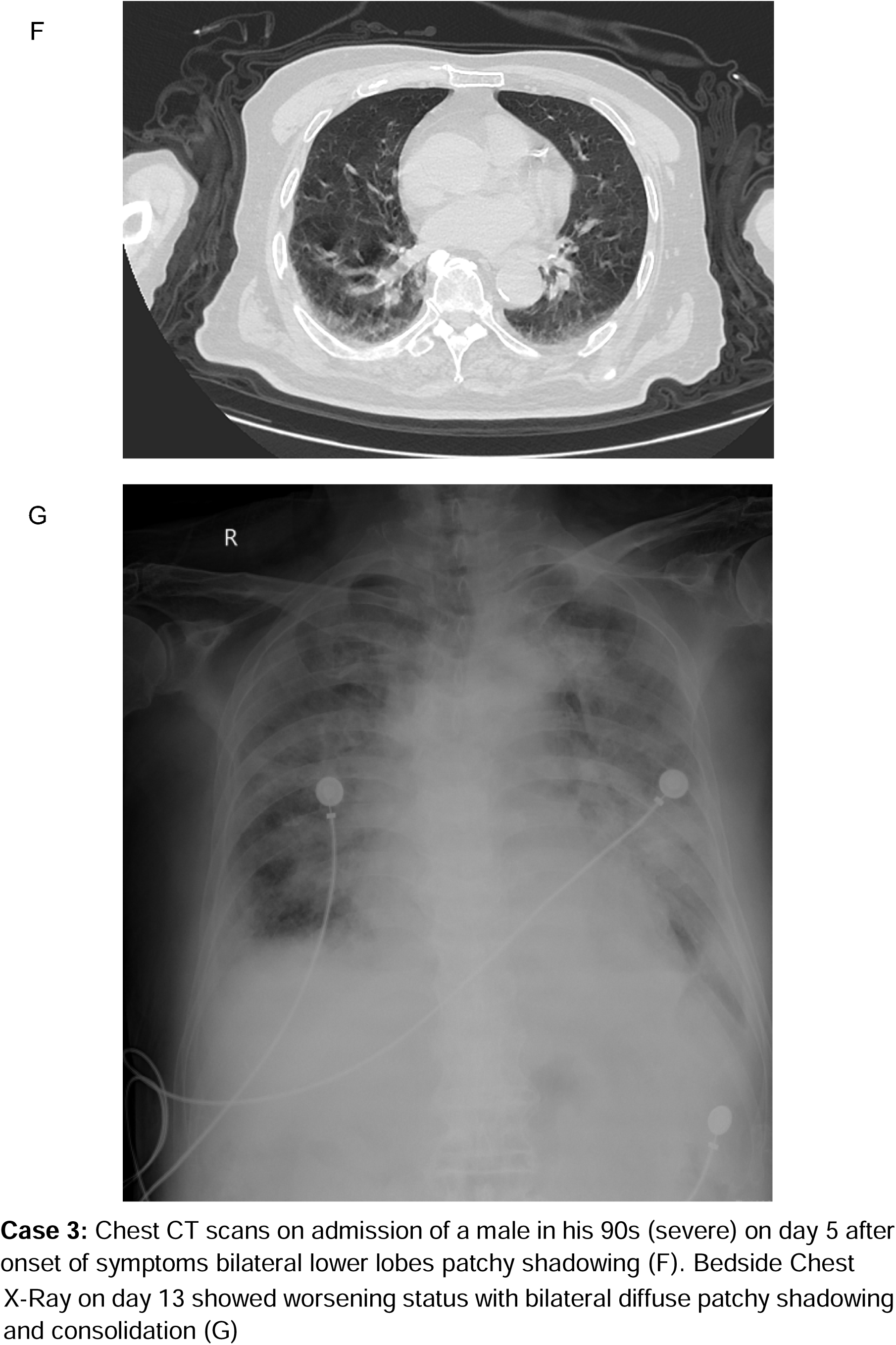
Representative chest radiographic manifestations in 3 patients with Omicron variant

### Viral eradication in the upper respiratory tract

All patients had daily PCR tested for Omicron variant in upper respiratory tract samples during this observation period. The median interval of PCR negative conversion was 10 days (IQR: 8-13 days) in all patients. In four asymptomatic patients, it took 9 days for the viral RNA converting to be negative after admission(IQR: 8–10.5 days). The median time of PCR conversation was significantly longer in moderate patients than those in mild (13 days, IQR: 10-15 days, vs. 10 days, IQR:8-12.5 days (p=0.031)(**Fig.6**). The PCR conversion time in patient under 14 was significantly shorter than patients above 14 (8 days, IQR: 7-9 days vs. 11 days, IQR: 9-14days, p <0.0001) (**Fig.7**)

**Fig. 6.**
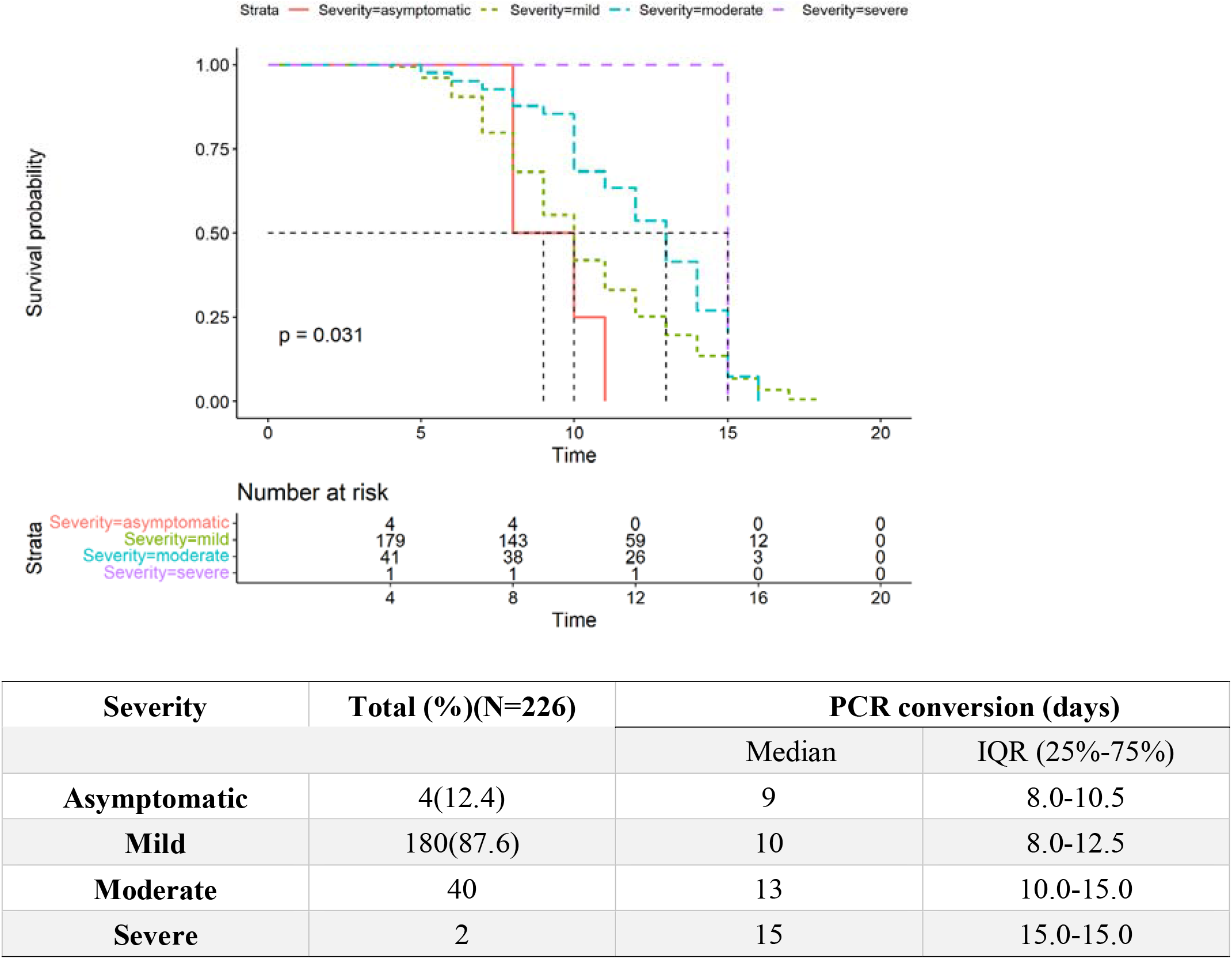
Time to a negative conversion of Omicron infection by PCR of upper respiratory tract samples among asymptomatic, mild, moderate and severe cases.

**Fig. 7.**
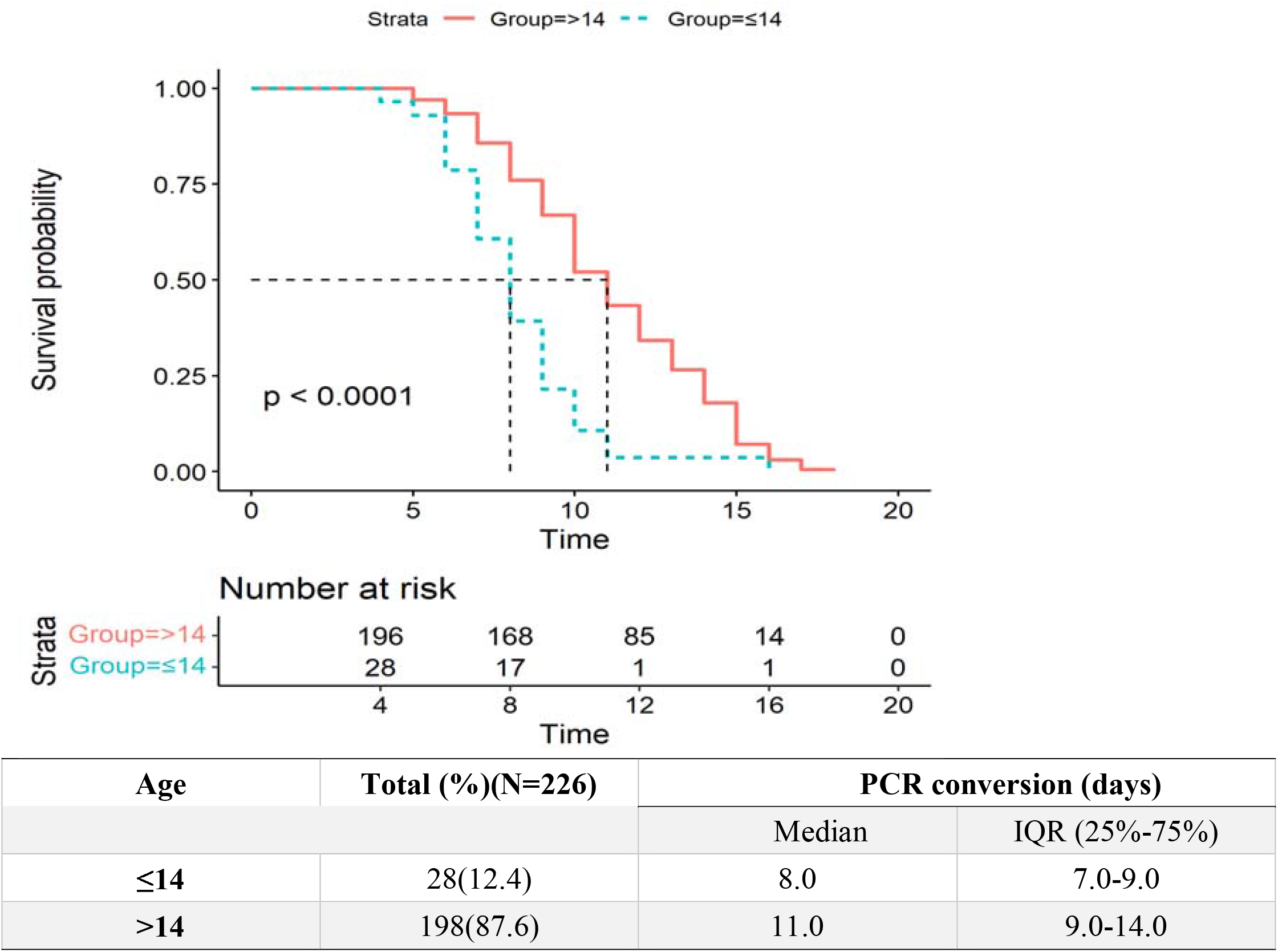
Time to a negative conversion of Omicron infection by PCR in patients under 14 and above 14

### Factors related to the severity of the infection

To figure out factors that were associated with the severity of the disease, we then compared clinical laboratory characters and vaccination status of asymptomatic(N = 4), mild (N = 180) and moderate (N = 41) patients.

In univariate analysis, older age, comorbidity, lymphopenia, high levels of C-reactive protein (CRP), erythrocyte sedimentation rate (ESR), lactate, estimated glomerular filtration rate (e-GFR), low levels of albumin, and less dose of vaccination are all associated with the severity of the Omicron infection (**Table 2, Table3**). In multivariate logistic analysis, ESR(OR=1.05) was independently related to the severity of the infection.

**Table 2.**
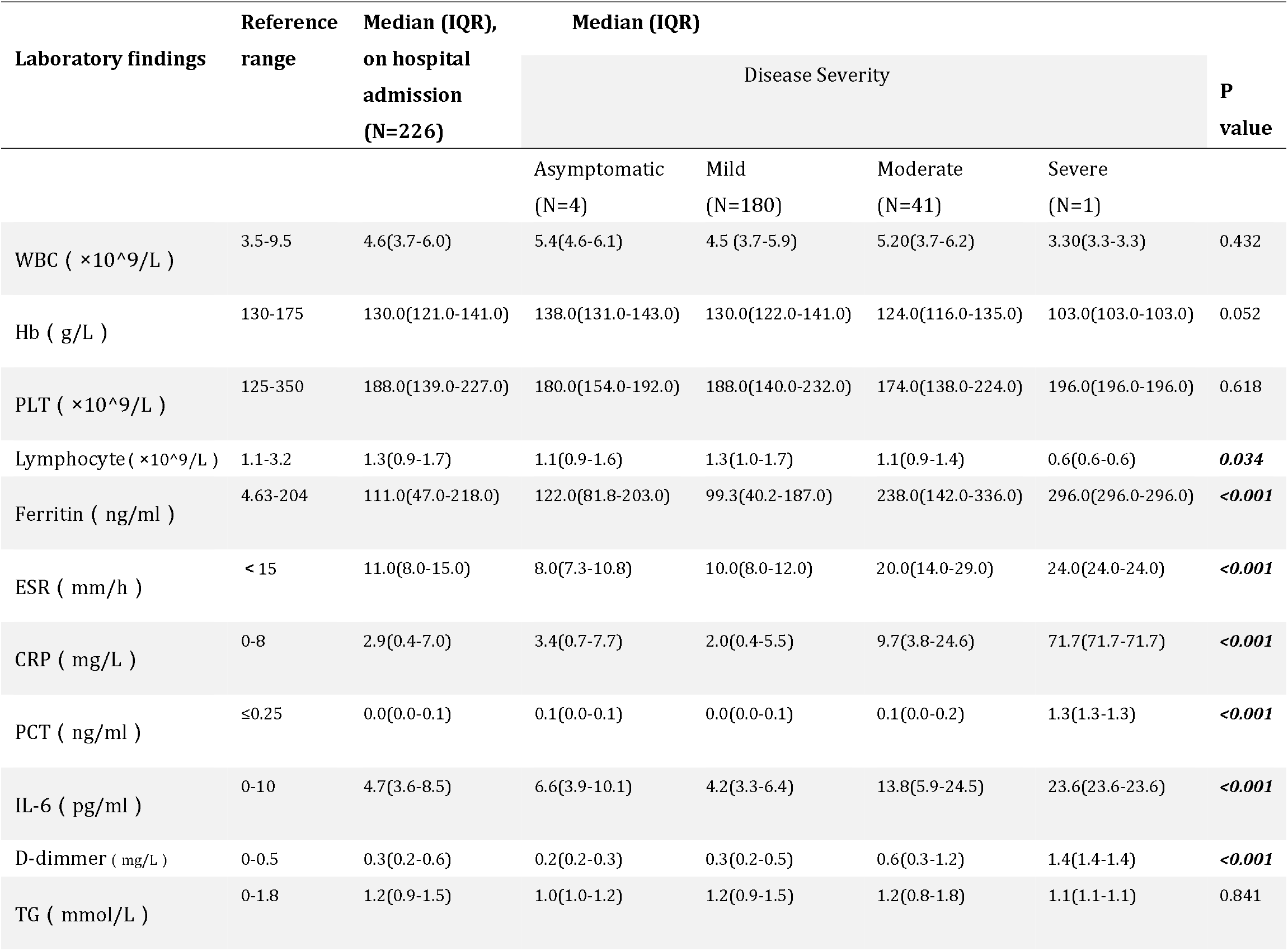
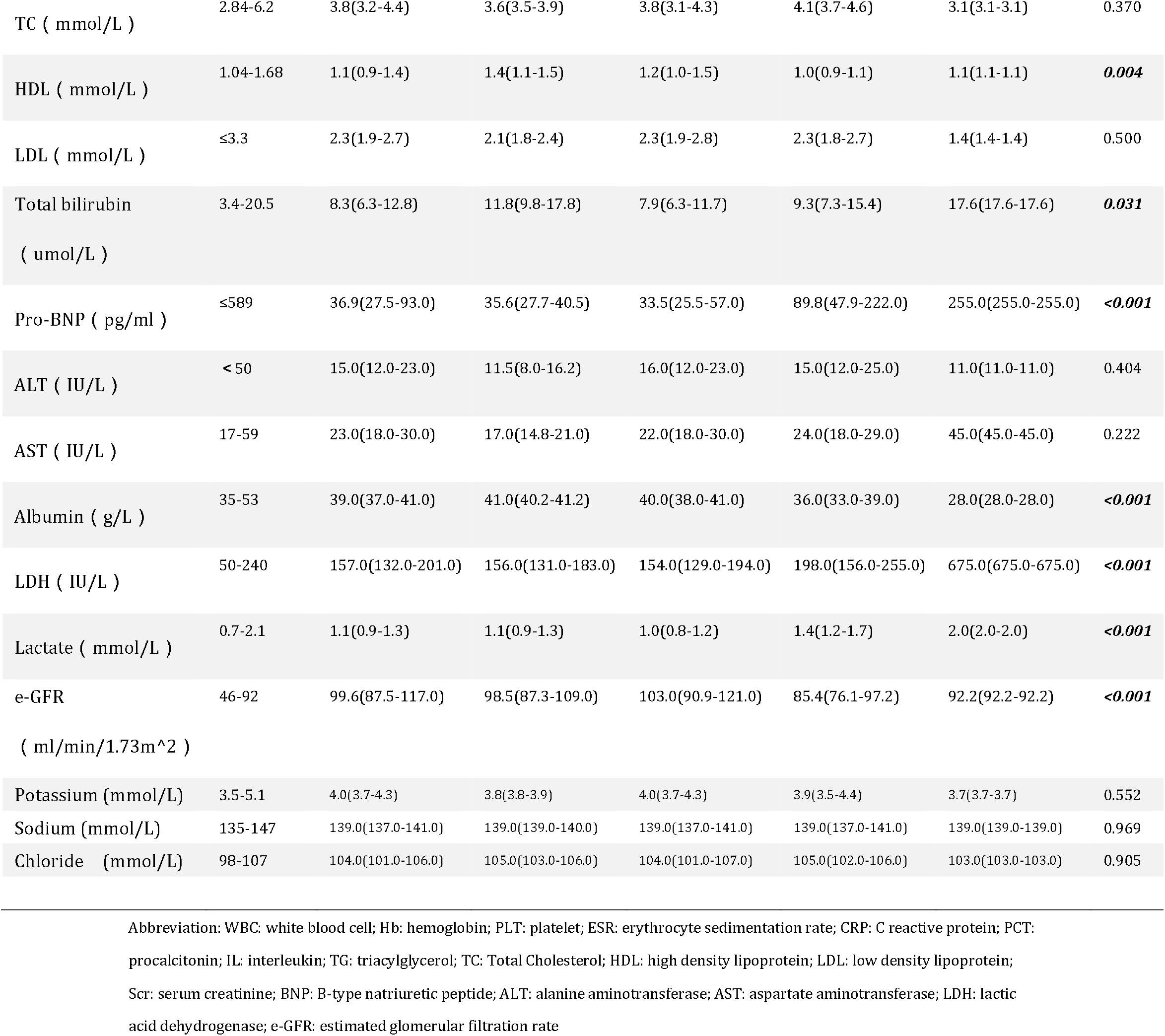
Laboratory findings on admission (N=226)

| Laboratory findings | Reference range | Median (IQR),<br>on hospital admission<br>(N=226) | Median (IQR) |  |  |  | P value |
| --- | --- | --- | --- | --- | --- | --- | --- |
|  |  |  | Disease Severity |  |  |  |  |
|  |  |  | Asymptomatic<br>(N=4) | Mild<br>(N=180) | Moderate<br>(N=41) | Severe<br>(N=1) |  |
| WBC ( ×10^9/L ) | 3.5-9.5 | 4.6(3.7-6.0) | 5.4(4.6-6.1) | 4.5 (3.7-5.9) | 5.20(3.7-6.2) | 3.30(3.3-3.3) | 0.432 |
| Hb ( g/L ) | 130-175 | 130.0(121.0-141.0) | 138.0(131.0-143.0) | 130.0(122.0-141.0) | 124.0(116.0-135.0) | 103.0(103.0-103.0) | 0.052 |
| PLT ( ×10^9/L ) | 125-350 | 188.0(139.0-227.0) | 180.0(154.0-192.0) | 188.0(140.0-232.0) | 174.0(138.0-224.0) | 196.0(196.0-196.0) | 0.618 |
| Lymphocyte( ×10^9/L ) | 1.1-3.2 | 1.3(0.9-1.7) | 1.1(0.9-1.6) | 1.3(1.0-1.7) | 1.1(0.9-1.4) | 0.6(0.6-0.6) | **0.034** |
| Ferritin ( ng/ml ) | 4.63-204 | 111.0(47.0-218.0) | 122.0(81.8-203.0) | 99.3(40.2-187.0) | 238.0(142.0-336.0) | 296.0(296.0-296.0) | **<0.001** |
| ESR ( mm/h ) | < 15 | 11.0(8.0-15.0) | 8.0(7.3-10.8) | 10.0(8.0-12.0) | 20.0(14.0-29.0) | 24.0(24.0-24.0) | **<0.001** |
| CRP ( mg/L ) | 0-8 | 2.9(0.4-7.0) | 3.4(0.7-7.7) | 2.0(0.4-5.5) | 9.7(3.8-24.6) | 71.7(71.7-71.7) | **<0.001** |
| PCT ( ng/ml ) | ≤0.25 | 0.0(0.0-0.1) | 0.1(0.0-0.1) | 0.0(0.0-0.1) | 0.1(0.0-0.2) | 1.3(1.3-1.3) | **<0.001** |
| IL-6 ( pg/ml ) | 0-10 | 4.7(3.6-8.5) | 6.6(3.9-10.1) | 4.2(3.3-6.4) | 13.8(5.9-24.5) | 23.6(23.6-23.6) | **<0.001** |
| D-dimmer ( mg/L ) | 0-0.5 | 0.3(0.2-0.6) | 0.2(0.2-0.3) | 0.3(0.2-0.5) | 0.6(0.3-1.2) | 1.4(1.4-1.4) | **<0.001** |
| TG ( mmol/L ) | 0-1.8 | 1.2(0.9-1.5) | 1.0(1.0-1.2) | 1.2(0.9-1.5) | 1.2(0.8-1.8) | 1.1(1.1-1.1) | 0.841 |
| TC ( mmol/L ) | 2.84-6.2 | 3.8(3.2-4.4) | 3.6(3.5-3.9) | 3.8(3.1-4.3) | 4.1(3.7-4.6) | 3.1(3.1-3.1) | 0.370 |
| HDL ( mmol/L ) | 1.04-1.68 | 1.1(0.9-1.4) | 1.4(1.1-1.5) | 1.2(1.0-1.5) | 1.0(0.9-1.1) | 1.1(1.1-1.1) | **0.004** |
| LDL ( mmol/L ) | ≤3.3 | 2.3(1.9-2.7) | 2.1(1.8-2.4) | 2.3(1.9-2.8) | 2.3(1.8-2.7) | 1.4(1.4-1.4) | 0.500 |
| Total bilirubin<br>( umol/L ) | 3.4-20.5 | 8.3(6.3-12.8) | 11.8(9.8-17.8) | 7.9(6.3-11.7) | 9.3(7.3-15.4) | 17.6(17.6-17.6) | **0.031** |
| Pro-BNP ( pg/ml ) | ≤589 | 36.9(27.5-93.0) | 35.6(27.7-40.5) | 33.5(25.5-57.0) | 89.8(47.9-222.0) | 255.0(255.0-255.0) | **<0.001** |
| ALT ( IU/L ) | < 50 | 15.0(12.0-23.0) | 11.5(8.0-16.2) | 16.0(12.0-23.0) | 15.0(12.0-25.0) | 11.0(11.0-11.0) | 0.404 |
| AST ( IU/L ) | 17-59 | 23.0(18.0-30.0) | 17.0(14.8-21.0) | 22.0(18.0-30.0) | 24.0(18.0-29.0) | 45.0(45.0-45.0) | 0.222 |
| Albumin ( g/L ) | 35-53 | 39.0(37.0-41.0) | 41.0(40.2-41.2) | 40.0(38.0-41.0) | 36.0(33.0-39.0) | 28.0(28.0-28.0) | **<0.001** |
| LDH ( IU/L ) | 50-240 | 157.0(132.0-201.0) | 156.0(131.0-183.0) | 154.0(129.0-194.0) | 198.0(156.0-255.0) | 675.0(675.0-675.0) | **<0.001** |
| Lactate ( mmol/L ) | 0.7-2.1 | 1.1(0.9-1.3) | 1.1(0.9-1.3) | 1.0(0.8-1.2) | 1.4(1.2-1.7) | 2.0(2.0-2.0) | **<0.001** |
| e-GFR<br>( ml/min/1.73m^2 ) | 46-92 | 99.6(87.5-117.0) | 98.5(87.3-109.0) | 103.0(90.9-121.0) | 85.4(76.1-97.2) | 92.2(92.2-92.2) | **<0.001** |
| Potassium (mmol/L) | 3.5-5.1 | 4.0(3.7-4.3) | 3.8(3.8-3.9) | 4.0(3.7-4.3) | 3.9(3.5-4.4) | 3.7(3.7-3.7) | 0.552 |
| Sodium (mmol/L) | 135-147 | 139.0(137.0-141.0) | 139.0(139.0-140.0) | 139.0(137.0-141.0) | 139.0(137.0-141.0) | 139.0(139.0-139.0) | 0.969 |
| Chloride (mmol/L) | 98-107 | 104.0(101.0-106.0) | 105.0(103.0-106.0) | 104.0(101.0-107.0) | 105.0(102.0-106.0) | 103.0(103.0-103.0) | 0.905 |

Abbreviation: WBC: white blood cell; Hb: hemoglobin; PLT: platelet; ESR: erythrocyte sedimentation rate; CRP: C reactive protein; PCT: procalcitonin; IL: interleukin; TG: triacylglycerol; TC: Total Cholesterol; HDL: high density lipoprotein; LDL: low density lipoprotein; Scr: serum creatinine; BNP: B-type natriuretic peptide; ALT: alanine aminotransferase; AST: aspartate aminotransferase; LDH: lactic acid dehydrogenase; e-GFR: estimated glomerular filtration rate

**Table 3.** Factors related to the severity of the Omicron infection.

|  | Univariate logistic regression |  |  | Multivariate logistic regression |  |  |
| --- | --- | --- | --- | --- | --- | --- |
|  | OR | 95%CI | P value | OR | 95%CI | P value |
| Age | 1.06 | 1.03-1.08 | **<0.001** | 1.03 | 0.99-1.08 | 0.138 |
| Female | 0.58 | 0.29-1.15 | 0.120 | 0.51 | 0.20-1.24 | 0.143 |
| Fever | 1.11 | 0.56-2.19 | 0.757 | 1.97 | 0.75-5.35 | 0.172 |
| Comorbidity | 5.46 | 2.46-12.12 | **<0.001** | 1.11 | 0.33-3.69 | 0.867 |
| WBC | 1.09 | 0.94-1.26 | 0.266 | 0.95 | 0.74-1.20 | 0.674 |
| Lymphocytes | 0.48 | 0.25-0.91 | **0.025** | 0.70 | 0.27-1.65 | 0.459 |
| CRP | 1.03 | 1.01-1.05 | **0.004** | 0.99 | 0.96-1.02 | 0.387 |
| ESR | 1.08 | 1.04-1.13 | **<0.001** | 1.05 | 1.02-1.10 | **0.007** |
| Albumin | 0.81 | 0.73-0.89 | **<0.001** | 0.95 | 0.84-1.07 | 0.363 |
| ALT | 1.00 | 0.99-1.01 | 0.861 | 1.00 | 0.97-1.04 | 0.908 |
| AST | 1.00 | 0.99-1.00 | 0.920 | 1.00 | 0.97-1.02 | 0.703 |
| Lactate | 6.51 | 2.72-15.58 | **<0.001** | 2.34 | 0.67-8.25 | 0.180 |
| e-GFR | 0.97 | 0.95-0.98 | **<0.001** | 0.99 | 0.97-1.02 | 0.606 |
| Dose of vaccinations |  |  |  |  |  |  |
| 0 | 1.00 |  |  | 1.00 |  |  |
| 1 | 1.46 | 0.25-8.49 | 0.676 | 2.43 | 0.17-28.85 | 0.492 |
| 2 | 0.63 | 0.28-1.46 | 0.282 | 0.71 | 0.21-2.25 | 0.571 |
| 3 | 0.28 | 0.11-0.73 | **0.009** | 0.52 | 0.15-1.73 | 0.296 |

## Discussion

On November 24, 2021, World Health Organization (WHO) announced a new variant of SARS-CoV-2 Omicron in South Africa, and 17days after, the first case infected with Omicron in China (11). Recent investigations on Omicron variant have generally been described with the epidemiologic characteristics, initial clinical, laboratory and radiological findings(4, 6, 12). To our knowledge, we are the first to describe the temporal clinical advancement of Omicron variant infection to date. Our investigation has several distinguished features from current research. As one of the megacities and most important economic centers in China, the policy of “Dynamic zero” was implemented strictly in Shanghai. The whole city underwent completely lockdown during the pandemic outbreak since the end of March 2022(13). Under the policy of “all those in need have been tested, and if positive, have been quarantined, hospitalized, or treated” in China, 25 million residents had been daily tested for the Omicron strain infection by RT-PCR. All positive patients without symptoms were transferred to mobile cabin hospitals. Those symptomatic patients who have potential to exacerbate were directly sent to the fever clinic or COVID-19 designated hospital. All participants in our study came from mobile cabin hospitals, fever clinics and nursing homes which can be inclusive and comprehensive, and representative of the current real -world population.

In this study, most individuals infected by omicron were mild or moderate and did not aggravate into severe or critical conditions. A recent investigation showed Omicron can cause lower severity disease than the Delta variant, which may be attributed to Omicron replicates less efficiently in the lung and more efficiently in the respiratory tract(14). However, it has high transmissibility, which lead to nearly 0.62 million Shanghai people being infected in 2 months. To prevent and control the Omicron infection and its related diseases, exploring the clinical progression and factors associated with the severity of the infection is needed.

In accordance with a previous study from the U.S. CDC (6), we found that as high as 74.3% of the patients with the mutant were suffering from cough, which indicated that the main attacking site of the virus is located on the upper airways(15). Coughing can generate viral particles contained aerosols which can be expelled easily from the nose and mouth. This could explain why the variant can spread so fast in such a short period of time(16). An investigation confirmed that covering your mouth can speed the SARS-CoV-2 RNA PCR conversion in patients when they contracted the virus (17). Therefore, wearing a face mask is strongly recommended for stopping the higher transmissibility of the Omicron variant.

The infectiousness and transmissibility of SARS-CoV-2 has been associated with viral shedding(18, 19). To date, there are few studies on the predictors of Omicron variant shedding duration. An interpretation for the Omicron variant eradication in the upper respiratory tract is very important for the implementation of preventive tactics such as determination of isolation period. Data from Japan suggest that the amount of viral RNA reached its highest three to six days after diagnosis or symptom emergence(20). The median time after onset of symptoms to viral clearance in this study was 10 days, which was close to that in Covid-19(21), but significantly shorter than that of MERS (17 days)(22). In addition, no radiographic progressions were identified in 98% of patients after hospitalization, which could be attributed to its lower virulence and having less chance to cause havoc in the lungs(23, 24).

It was reported that administration of Paxlovid made contributions to reduce hospital admissions and deaths among individuals with high-risk, especially for elderly. In our research, Paxlovid was also proved to significantly reduced the period of viral PCR conversion in patients whose conditions were mild and moderate. As a traditional Chinese medicine, LHQW can significantly suppress the SARS-COV-2 replication affecting the morphology of viral granule and exerting antiviral activity in vitro(25, 26). Some studies showed that it may improve the clinical symptoms of patients infected with SARS-COV-2, such as fever, fatigue and myalgia, but safety is not guaranteed if combined with conventional treatment(27). In this retrospective study, it did not shorten the interval of PCR conversion in mild cases aged above 14 compared with the Paxlovid-treated group. However, it probably can alleviate the clinical symptoms of Omicron variant infection including cough, expectoration and rhinorrhea.

Although cough is the most common symptom in Omicron infection, fever is still an important indicator for uncontrolled viral replication in the body. The median duration of fever in all febrile patients was 5 days, which was significantly shorter than that in MERS (8 days), Covid-19 (10 days) and SARS (11.4 days), indicating its decreased virulence (21).

Covid-19 inactivated vaccine was the main type of vaccine used in China’s national inoculation program. Currently, 95.1% and 42.5% of Shanghai permanent residents received full and boosted vaccination, respectively. Only 39.3% of people aged above 60 got boosted vaccination (8). Our study manifested that there was a significant difference regarding severity between patients who received booster vaccinations and who were unvaccinated, partially vaccinated or fully vaccinated. This result is consistent with research from Hongkong which confirmed that both boosted vaccination of inactivated and mRNA vaccines can provide sufficient protective capability against severe/fatal cases for all Omicron-infected individuals(28). This phenomenon might be attributable to neutralizing antibodies elicited by the vaccines that all patients inoculated, The discovery was in line with another finding stating unvaccinated patients infected with the preceding variants of SARS-CoV-2 showed occasional neutralization to the omicron variant as well(29). As the virus continues to evolve, we cannot predict the subsequent mutating direction of the virus and whether its causes will be more or less severe. The whole society must constantly adapt by increasing their immunity through vaccination(30).

A recent investigation by Butt A et al. showed Omicron variant infection in children who did not have prior infection or vaccination are associated with less severe disease than the Delta variant infection(31). Our findings indicated the Omicron variant seemed to be ‘friendly’ to children with the PCR conversion interval of 8 days and mild symptoms even without any antiviral drugs. As already known, young children are especially vulnerable to have upper airway infection due to their relatively narrow and collapsible nasal passages, and that babies breathe only through their noses(32), which means the virus mainly lingers in the upper respiratory tract. But at the same time, the virus can be relatively driven out by breathing or coughing. Nevertheless, the actual mechanism behind this phenomenon needs to be elucidated in the coming study.

This study has several limitations. First, we did not test dynamic changes of IgM and IgG in Omicron infected patients, so we cannot determine what protective degree the vaccinated patients have. Second, we only tested viral PCR conversion in nasopharyngeal and not in sera, stool, or urine which also can affect viral shedding time (VST) (33, 34). However, a previous study has shown that the VST of SARS-CoV-2 in feces was longer than that in respiratory tract sample raising the concern that the virus could infect individuals by fecal-oral transmission (35). Third, two patients were still hospitalized at the time of manuscript submission, therefore, clinical outcomes of the patients can’t be obtained and continued follow-up is still needed.

In conclusion, the majority of the Omicron variant cases in this study are not severe. The disease progression suggests that population-based booster vaccination and early control of viral replication by application of Paxlovid are essential to improve the prognosis of Omicron variant infection, especially in older populations and those who are suffering from coexisting illnesses.

## Data Availability

All data produced in the present study are available upon reasonable request to the authors

## Acknowledgments

The authors wish to thank all patients involved in the study and the following individuals for their help in data collection as well as their medical advices and consultations during taking care of all patients: Drs. Sunying Hua, Baoliang Shen, Peng Xu, Feipeng Xu, Ruixing Xian, Hongfang Tao, Lizhen Liu.

